# Type I Interferon Signature Strength Correlates with Alloimmunization-Associated Transcriptomic Programs in Systemic Lupus Erythematosus: A Multi-Cohort Analysis

**DOI:** 10.64898/2026.04.04.26350150

**Authors:** Jaeeun Yoo

**Affiliations:** Department of Laboratory Medicine, Incheon St. Mary’s Hospital, College of Medicine, The Catholic University of Korea, Incheon, South Korea

**Author notes:** Corresponding author: Jaeeun Yoo, M.D., Department of Laboratory Medicine, Incheon St. Mary’s Hospital, College of Medicine, The Catholic University of Korea, Incheon, South Korea.

**Keywords:** systemic lupus erythematosus, type I interferon, RBC alloimmunization, transcriptomics, RNA-seq, interferon signature, transfusion immunology, gene set enrichment analysis

## Abstract

Red blood cell (RBC) alloimmunization is a clinically significant complication in transfused patients whose immunological determinants remain incompletely understood. Type I interferon (IFN-I) signaling drives RBC alloimmunization in murine models, and systemic lupus erythematosus (SLE) is characterized by constitutive IFN-I hyperactivation alongside elevated alloimmunization rates. We analyzed three publicly available SLE RNA-seq cohorts (GSE72509, GSE112087, GSE122459; whole blood and PBMC; total n = 150 SLE) in a pre-specified discovery–replication–validation design. A 14-gene IFN-I signature score was computed per sample; differential expression, gene set enrichment analysis, and Spearman correlation were performed independently per cohort. IFN-I scores were significantly elevated in SLE versus healthy controls in all three cohorts (p < 0.01 each). IFN-high SLE patients showed 665 differentially expressed genes, with enrichment of alloimmunization-associated and plasmablast differentiation gene sets confirmed by GSEA. The alloimmunization signature score correlated significantly with IFN-I score across all three independent cohorts (ρ = +0.77, +0.51, +0.60; all FDR q < 0.05); Tfh differentiation showed no association in any cohort. These findings suggest that IFN-I pathway activity in SLE is associated with an alloimmunization-prone transcriptomic landscape, To our knowledge, this represents the first human transcriptomic evidence that IFN-I pathway activity in SLE is coupled to alloimmunization-associated immune programs in vivo. These findings identify IFN-I score as a candidate biomarker of alloimmunization susceptibility in SLE and provide translational rationale for prospective studies incorporating transfusion outcome data.

## Introduction

Red blood cell (RBC) alloimmunization—the development of antibodies against donor RBC antigens following transfusion—is a clinically significant complication in patients requiring repeated transfusions^1^. Once alloantibodies form, they complicate future crossmatching, restrict the pool of compatible blood units, and increase the risk of acute and delayed hemolytic transfusion reactions^1^. Despite its clinical importance, the immunological mechanisms that predispose certain patients to alloimmunization remain incompletely understood.

Type I interferon (IFN-I) signaling has emerged as a key immunological driver of RBC alloimmunization. In murine transfusion models, IFN-I signaling through the IFNα/β receptor (IFNAR) has been shown to be necessary and sufficient for inflammation-induced RBC alloimmunization^2^. Supporting this, in vitro evidence suggests that IFN-I can broadly promote the humoral immune programs underlying alloantibody generation^3^. The IFN-I transcriptomic signature, measured by coordinated upregulation of interferon-stimulated genes (ISGs) in peripheral blood, has been validated as a reliable index of IFN-I pathway activity across patient cohorts^4,5^.

Systemic lupus erythematosus (SLE) is among the conditions most strongly characterized by IFN-I pathway hyperactivation, with an elevated IFN-I transcriptomic signature present in 60–80% of patients and closely linked to disease activity^4,5,6^. SLE patients also frequently develop cytopenias—including anemia of chronic disease and autoimmune hemolytic anemia—that necessitate repeated RBC transfusion^7,8^. Notably, patients with chronic inflammatory autoimmune diseases, including SLE, develop RBC alloantibodies at substantially higher rates than the general transfused population, underscoring a disease-specific immunological susceptibility that remains mechanistically unresolved^1,8^.

Yet whether the degree of IFN-I pathway activation in SLE is transcriptomically coupled to alloimmunization-associated immune programs in vivo has not been directly examined in human patients. This represents a critical translational gap: if IFN-I signature strength predicts alloimmunization-associated immune programs, IFN-I score could serve as a clinically accessible biomarker to identify SLE patients at elevated alloimmunization risk prior to transfusion—and therapies targeting the IFN-I pathway (such as the approved anti-IFNAR antibody anifrolumab) could potentially modulate this risk. We therefore analyzed publicly available RNA-seq datasets across three independent SLE cohorts to determine, for the first time in human transcriptomic data, whether IFN-I signature strength correlates with gene expression programs relevant to alloimmunization risk.

## Materials and Methods

### Study Design

We employed a pre-specified three-cohort discovery–replication–validation design (Figure 1). GSE72509 (n = 99 SLE, 18 HC; whole blood RNA-seq; Hung et al.) served as the discovery cohort. GSE112087 (n = 31 SLE, 29 HC; whole blood RNA-seq; Figgett et al.) served as the whole-blood replication cohort. GSE122459 (n = 20 SLE, 6 HC; PBMC RNA-seq; Tokuyama et al.) provided cross-platform validation, allowing assessment of whether findings generalize across sample types. All three cohorts are publicly available from NCBI GEO. All cohorts were analyzed entirely independently with no cross-dataset pooling or batch correction. Cross-cohort replication was defined a priori as nominal p < 0.05 in the same direction in at least two of three independent cohorts. As this study constitutes a re-analysis of publicly available, de-identified data, no institutional ethics approval was required.

**Figure 1.**
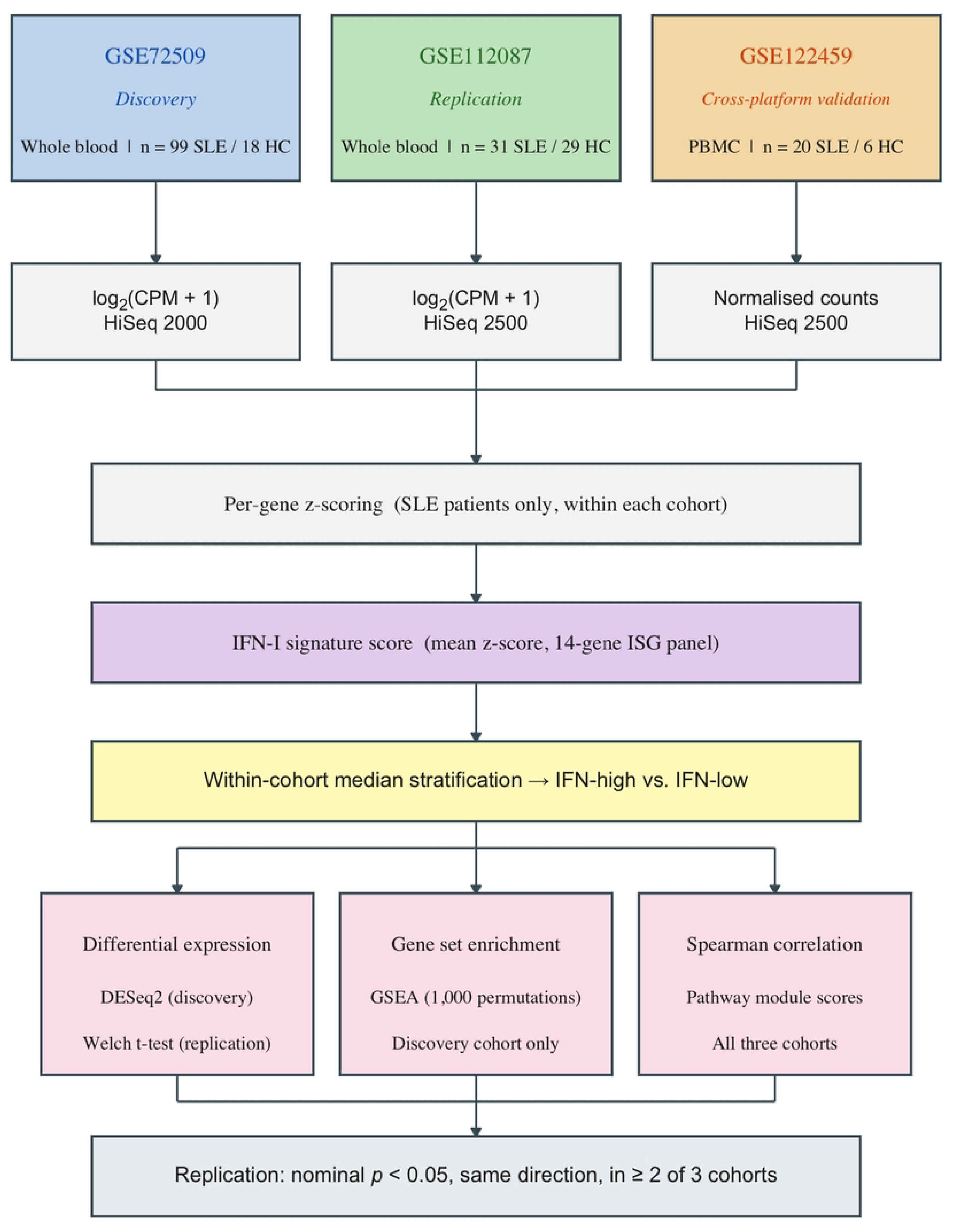
Study design flowchart. Three independent SLE transcriptomic cohorts were analyzed in a pre-specified discovery–replication–cross-platform validation design. GSE72509 (discovery, PAXgene whole blood, n = 99 SLE / 18 HC) and GSE112087 (replication, whole blood, n = 31 SLE / 29 HC) share the same sample type (whole blood, RNA-seq). GSE122459 (cross-platform validation, PBMC, n = 20 SLE / 6 HC) assessed generalizability across sample types. Each cohort underwent independent CPM normalization (GSE112087) or direct value use, per-gene z-scoring, and IFN-I signature scoring. SLE patients were stratified by within-cohort median IFN-I score into IFN-high and IFN-low subgroups for all downstream analyses. HC, healthy control; WB, whole blood; PBMC, peripheral blood mononuclear cells.^9,10,11^

### Data Processing and IFN-I Signature Scoring

For GSE112087, raw counts were library-size normalized to counts per million (CPM) and log_2_(CPM + 1) transformed; pre-normalized expression values were used directly for GSE72509 and GSE122459. Within each dataset, all samples (SLE and healthy controls) were included in per-gene z-scoring (mean subtraction, unit-variance scaling across all samples in the cohort). This step centers expression relative to each cohort’s own baseline, removes platform-driven scale differences, and ensures that IFN-I signature scores and pathway module scores reflect within-cohort relative expression rather than absolute values, enabling interpretable cross-pathway comparisons within each independent cohort.

A 14-gene IFN-I signature score was computed per sample as the arithmetic mean of per-gene z-scores, using a previously validated panel of established IFN-stimulated genes4,11: *MX1, MX2, IFI44, IFI44L, IFIT1, IFIT3, ISG15, OAS1, OAS2, OAS3, RSAD2, SIGLEC1, HERC5*, and *CMPK2*. All 14 ISGs were present in all three datasets. Within each cohort independently, SLE patients were stratified by median IFN-I score: those with scores strictly above the cohort median were assigned to IFN-high and those at or below the median to IFN-low. For cohorts with an odd number of SLE patients (GSE72509, n = 99; GSE112087, n = 31), this yields slightly unequal group sizes (GSE72509: IFN-high n = 49, IFN-low n = 50; GSE112087: IFN-high n = 15, IFN-low n = 16). IFN-I score differences between SLE and healthy controls were assessed by two-sided Mann–Whitney U test.

### Discovery Cohort Analyses: Differential Expression and GSEA

Differential expression between IFN-high (n = 49) and IFN-low (n = 50) SLE patients was performed in the discovery cohort only (n = 99 SLE provides adequate statistical power for genome-wide FDR-controlled analysis; the replication and validation cohorts [n = 31 and n = 20] were underpowered for this purpose), using the Welch two-sample t-test on per-gene z-scores across all 26,367 expressed genes. Multiple testing correction used the Benjamini–Hochberg false discovery rate (FDR) procedure^12^. DEGs were defined as FDR < 0.05 and |log_2_FC| > 0.5. Genes were ranked for GSEA by sign(log_2_FC) × [−log_10_(FDR)].

Preranked GSEA was performed using the method of Subramanian et al.^13^ implemented in gseapy (v0.10.x; 1,000 permutations; seed = 42). Six gene sets were evaluated: ISG core panel (14 genes), alloimmunization-associated signature (curated from published alloantigen and murine alloimmunization literature^2^), complement activation, plasmablast differentiation, B cell activation, and Tfh differentiation. Significance threshold: FDR q < 0.05.

### Pathway Module Scoring and Spearman Correlation

Pathway module scores were computed per SLE patient as the arithmetic mean of per-gene z-scores for all gene set members present in that dataset. Spearman rank correlation (ρ) assessed associations between IFN-I score and each pathway score; 95% confidence intervals were estimated by 1,000-iteration bootstrap resampling. Benjamini–Hochberg FDR correction was applied across all pathways within each cohort. All analyses were performed independently per cohort.

Robustness check: Spearman correlations in GSE72509 were repeated using SLE-only z-scores and without z-scoring; the IFN-I–alloimmunization correlation remained strong under both alternatives (ρ = 0.83 and 0.86, respectively; both p < 0.001).

Sensitivity analysis: To confirm the IFN–alloimmunization association was not driven by shared IFN-regulatory gene membership, eight IFN-regulatory members of the alloimmunization set (*IRF7, IFIH1, DDX58, TLR7, MYD88, TRIM25, TICAM1, IFNAR1*) were excluded, retaining a 12-gene non-IFN-adjacent subset (*FCGR1A, FCGR2A, FCGR2B, FCGR3A, CR2, LILRB1, LILRB3, C1QC, C1QB, CD38, MZB1, TXNDC5*). Spearman correlation with IFN-I score was computed independently in all three cohorts.

### Statistical Analysis

All statistical analyses were performed in Python (version 3.10; Python Software Foundation, https://www.python.org). Differential expression was assessed by Welch two-sample t-test with Benjamini–Hochberg false discovery rate (FDR) correction. Gene set enrichment analysis was performed using the gseapy package (v0.10) with 1,000 permutations. Spearman rank correlation coefficients were calculated with 95% bootstrap confidence intervals (1,000 iterations). Two distinct significance frameworks were applied: (1) within-cohort inference used FDR q-values (Benjamini–Hochberg correction across all pathways within each cohort); (2) cross-cohort replication was defined a priori as nominal p < 0.05 in the same direction in at least two of three independent cohorts. Significance notation reflects within-cohort FDR q-values: *** q < 0.001; * q < 0.05; ns q ≥0.05.

## Results

### IFN-I Signature Score Is Significantly Elevated in SLE Across All Three Cohorts

Cohort characteristics are summarized in Table 1. IFN-I scores were significantly elevated in SLE patients compared to healthy controls in all three cohorts (GSE72509: median SLE 0.54 vs. HC −1.21, p = 8.9 × 10−7; GSE112087: median SLE 0.94 vs. HC −0.82, p = 1.2 × 10−5; GSE122459: median SLE 0.58 vs. HC −0.99, p = 5.5 × 10−3; Mann–Whitney U test; Figure 2). Median stratification yielded IFN-high and IFN-low subgroups within each cohort: GSE72509 IFN-high n = 49 vs. IFN-low n = 50; GSE112087 IFN-high n = 15 vs. IFN-low n = 16; GSE122459 n = 10 vs. 10 (Figure 2).

**Table 1.** Baseline Cohort Characteristics.

| Characteristic | GSE72509<sup>a</sup> (Hung et al., 2015) | GSE112087<sup>b</sup> (Figgett et al., 2019) | GSE122459<sup>c</sup> (Tokuyama et al., 2018) |
| --- | --- | --- | --- |
| Dataset Overview |  |  |  |
| GEO accession | GSE72509 | GSE112087 | GSE122459 |
| Sample type | Whole blood | Whole blood | PBMC |
| Sequencing platform | Illumina HiSeq 2000 | Illumina HiSeq 2500 | Illumina HiSeq 2500 / NextSeq 500 |
| SLE patients, n | 99 | 31 | 20 |
| Healthy controls, n | 18 | 29 | 6 |
| Disease status | Active SLE | Active SLE | Active SLE |
| Country/Institution | USA (UCSD/Genentech) | Australia (Monash Medical Centre) | USA (Yale) |
| IFN Signature Groups (This Analysis) |  |  |  |
| IFN-high (strictly above median IFN-I score), n | 49 | 15 | 10 |
| IFN-low (at or below median IFN-I score), n | 50 | 16 | 10 |

<sup>a</sup> GSE72509: PAXgene whole blood RNA-seq from SLE patients and healthy controls enrolled in the ROSE Phase II clinical trial (Kalunian et al., Ann Rheum Dis 2016;75:196–202; Hung et al., Science 2015;350(6259):455–459).
<sup>b</sup> GSE112087: whole blood collected in PAXgene tubes from SLE patients and healthy donors at Monash Medical Centre, Melbourne, Australia (Figgett et al., Clin Transl Immunology 2019;8:e01093).
<sup>c</sup> GSE122459: PBMC collected from SLE patients and healthy donors (Tokuyama et al., Proc Natl Acad Sci USA 2018;115(50):12565–12572).
<sup>d</sup> IFN-high and IFN-low groups defined by median split of IFN-I signature scores within each dataset independently. IFN-I signature score = mean z-score of 14 ISGs: *MX1*, *MX2*, *IFI44*, *IFI44L*, *IFIT1*, *IFIT3*, *ISG15*, *OAS1*, *OAS2*, *OAS3*, *RSAD2*, *SIGLEC1*, *HERC5*, and *CMPK2*.
IFN-I, type I interferon; PBMC, peripheral blood mononuclear cells; SLE, systemic lupus erythematosus.

**Table 2.**
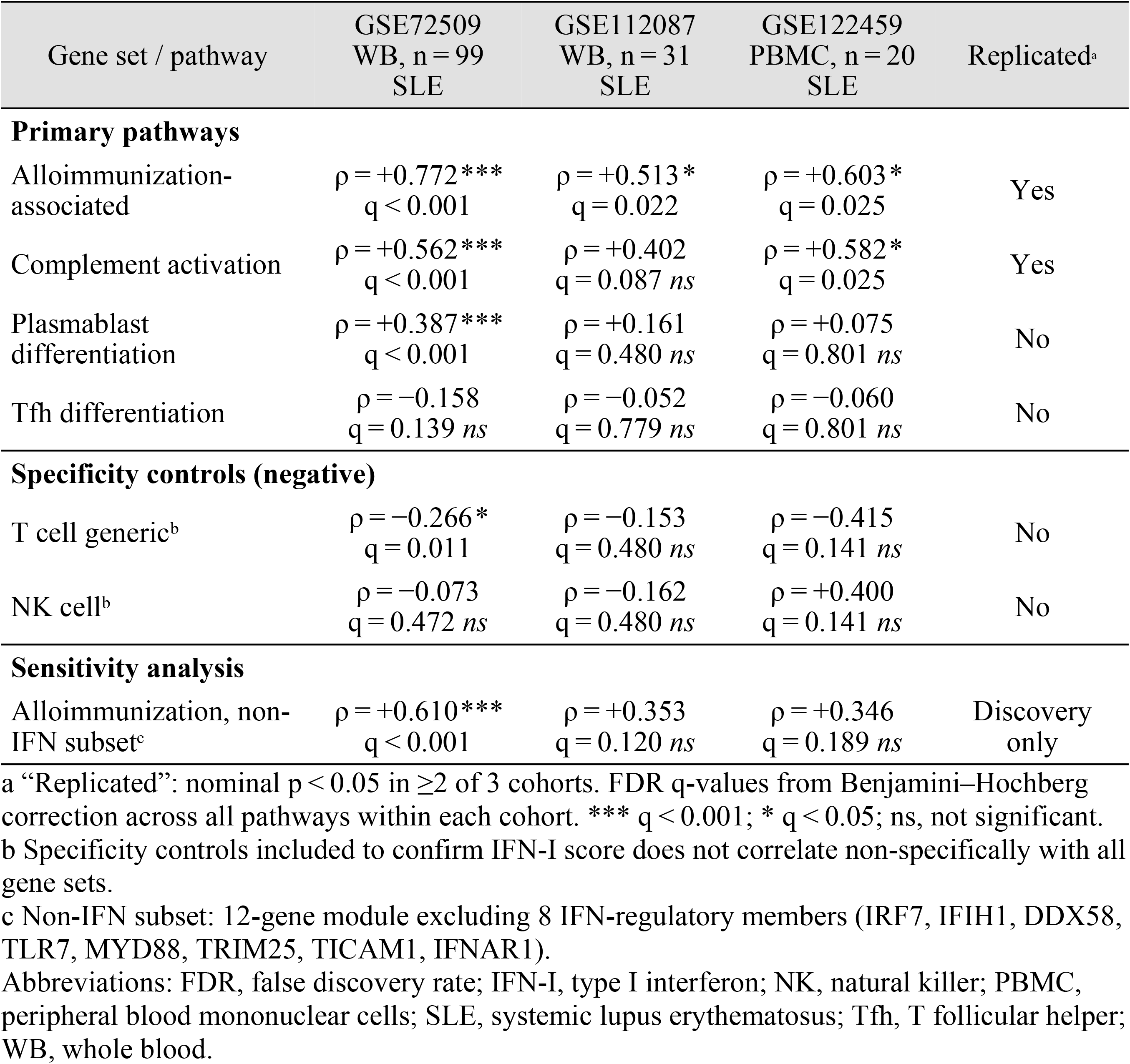
Spearman correlations between IFN-I signature score and pathway module scores across three independent SLE cohorts.

| Gene set / pathway | GSE72509<br>WB, n = 99<br>SLE | GSE112087<br>WB, n = 31<br>SLE | GSE122459<br>PBMC, n = 20<br>SLE | Replicated<sup>a</sup> |
| --- | --- | --- | --- | --- |
| **Primary pathways** |  |  |  |  |
| Alloimmunization-associated | $\rho = +0.772^{***}$<br>$q < 0.001$ | $\rho = +0.513^{*}$<br>$q = 0.022$ | $\rho = +0.603^{*}$<br>$q = 0.025$ | Yes |
| Complement activation | $\rho = +0.562^{***}$<br>$q < 0.001$ | $\rho = +0.402$<br>$q = 0.087$ ns | $\rho = +0.582^{*}$<br>$q = 0.025$ | Yes |
| Plasmablast differentiation | $\rho = +0.387^{***}$<br>$q < 0.001$ | $\rho = +0.161$<br>$q = 0.480$ ns | $\rho = +0.075$<br>$q = 0.801$ ns | No |
| Tfh differentiation | $\rho = -0.158$<br>$q = 0.139$ ns | $\rho = -0.052$<br>$q = 0.779$ ns | $\rho = -0.060$<br>$q = 0.801$ ns | No |
| **Specificity controls (negative)** |  |  |  |  |
| T cell generic<sup>b</sup> | $\rho = -0.266^{*}$<br>$q = 0.011$ | $\rho = -0.153$<br>$q = 0.480$ ns | $\rho = -0.415$<br>$q = 0.141$ ns | No |
| NK cell<sup>b</sup> | $\rho = -0.073$<br>$q = 0.472$ ns | $\rho = -0.162$<br>$q = 0.480$ ns | $\rho = +0.400$<br>$q = 0.141$ ns | No |
| **Sensitivity analysis** |  |  |  |  |
| Alloimmunization, non-IFN subset<sup>c</sup> | $\rho = +0.610^{***}$<br>$q < 0.001$ | $\rho = +0.353$<br>$q = 0.120$ ns | $\rho = +0.346$<br>$q = 0.189$ ns | Discovery only |

<sup>a</sup> “Replicated”: nominal $p < 0.05$ in $\geq 2$ of 3 cohorts. FDR q-values from Benjamini–Hochberg correction across all pathways within each cohort. \*\*\* $q < 0.001$ ; \* $q < 0.05$ ; ns, not significant.
<sup>b</sup> Specificity controls included to confirm IFN-I score does not correlate non-specifically with all gene sets.
<sup>c</sup> Non-IFN subset: 12-gene module excluding 8 IFN-regulatory members (IRF7, IFIH1, DDX58, TLR7, MYD88, TRIM25, TICAM1, IFNAR1).
Abbreviations: FDR, false discovery rate; IFN-I, type I interferon; NK, natural killer; PBMC, peripheral blood mononuclear cells; SLE, systemic lupus erythematosus; Tfh, T follicular helper; WB, whole blood.

**Figure 2.**
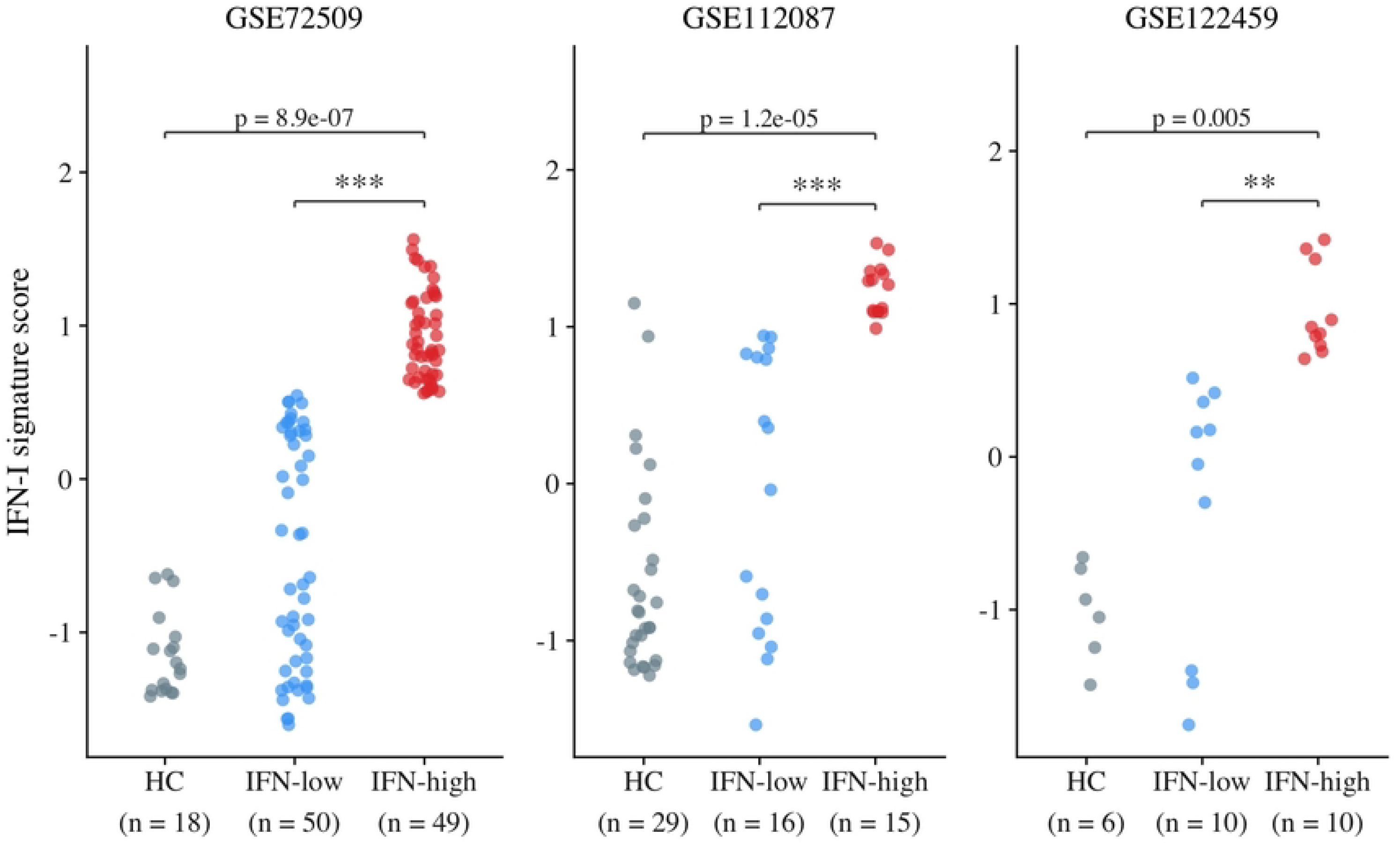
IFN-I signature score distribution across SLE patient subgroups and healthy controls in three independent cohorts. IFN-I score (mean z-score of 14 ISGs) is plotted for HC, IFN-low SLE, and IFN-high SLE subgroups separately for each cohort (left to right: GSE72509, GSE112087, GSE122459). IFN-high and IFN-low subgroups were defined by within-cohort median split of the IFN-I score. Each point represents one individual; horizontal bar indicates the group median. Significance brackets show t-test p-values for IFN-low vs. IFN-high (within SLE) and Mann–Whitney U p-values for HC vs. combined SLE. *p < 0.05; **p < 0.01; ***p < 0.001. HC, healthy control; ISG, interferon-stimulated gene; IFN-I, type I interferon; SLE, systemic lupus erythematosus.

### IFN-High SLE Patients Show Gene- and Pathway-Level Enrichment of Alloimmunization Programs

Differential expression analysis in the discovery cohort (IFN-high n = 49 vs. IFN-low n = 50) identified 665 DEGs (FDR < 0.05, |log_2_FC| > 0.5), with 538 genes upregulated and 127 downregulated in IFN-high patients (Figure 3; Supplementary Table S1). All 14 ISG panel members were among the most significantly upregulated genes (log_2_FC range +1.40 to +3.88; FDR < 10^−12^ for all 14), validating the median-split stratification approach. Among pathway-relevant upregulated DEGs, complement components (*SERPING1* [log_2_FC +2.28], *C2* [log_2_FC +1.01], *C1QC* [log_2_FC +0.99], *C1QB* [log_2_FC +0.76]; all FDR < 0.01), plasmablast markers (*MZB1* [log_2_FC +1.18, FDR = 0.001], *TXNDC5* [log_2_FC +0.87, FDR = 0.002], *CD38* [log_2_FC +1.09, FDR < 0.001]), and alloimmunization-associated innate immune sensors (*IRF7* [log_2_FC +1.86], *IFIH1* [log_2_FC +1.78], *DDX58* [log_2_FC +1.55], *MYD88* [log_2_FC +0.59]; all FDR < 0.001) were significantly upregulated in IFN-high patients. Among downregulated genes, the Tfh marker *CXCR5* (log_2_FC −0.54, FDR = 0.009) was notably reduced in IFN-high patients (Figure 3; Supplementary Table S1).

**Figure 3.**
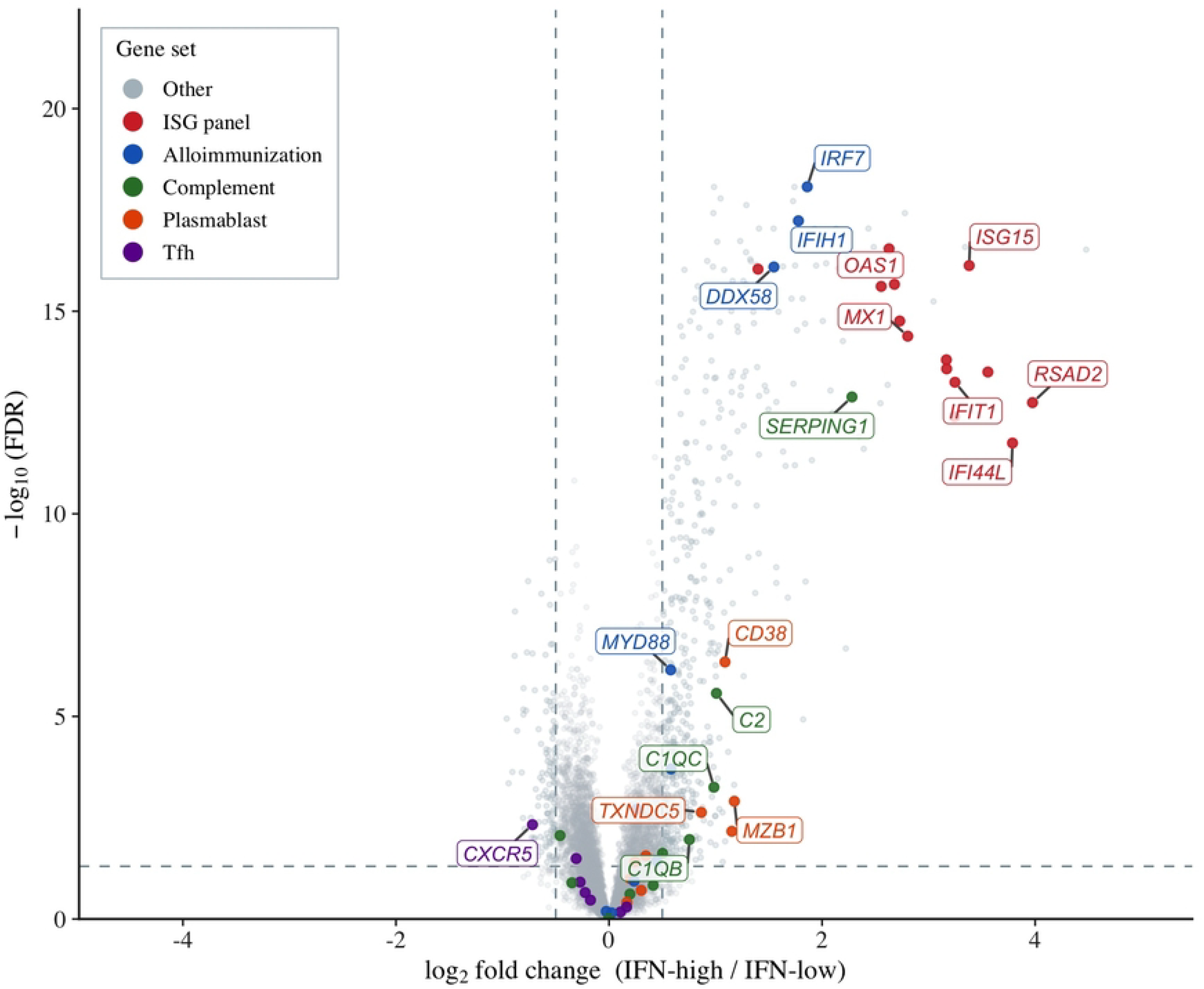
Volcano plot of differentially expressed genes between IFN-high (n = 49) and IFN-low (n = 50) SLE patients in the discovery cohort (GSE72509). The x-axis shows log_2_ fold change (IFN-high vs. IFN-low); the y-axis shows −log_10_(FDR-adjusted p-value). Dashed lines indicate |log_2_FC| = 0.5 and FDR = 0.05. Points are color-coded by gene set membership (legend inset). The ISG cluster (red) is shown enlarged in the zoom inset (upper right). Total: 538 upregulated and 127 downregulated genes meeting thresholds. Welch t-test with Benjamini– Hochberg FDR correction.

Preranked GSEA confirmed significant enrichment of the ISG core panel (NES = +2.70, FDR < 0.001), alloimmunization-associated gene set (NES = +2.34, FDR < 0.001), and plasmablast differentiation (NES = +1.94, FDR = 0.002) in IFN-high patients (Figure 4). Complement activation showed a trend toward enrichment that did not reach significance after FDR correction (NES = +1.52, FDR = 0.066). Tfh differentiation was not enriched (NES =− 1.09, FDR = 0.334) and B cell activation did not reach significance (NES = +0.82, FDR = 0.691).

**Figure 4.**
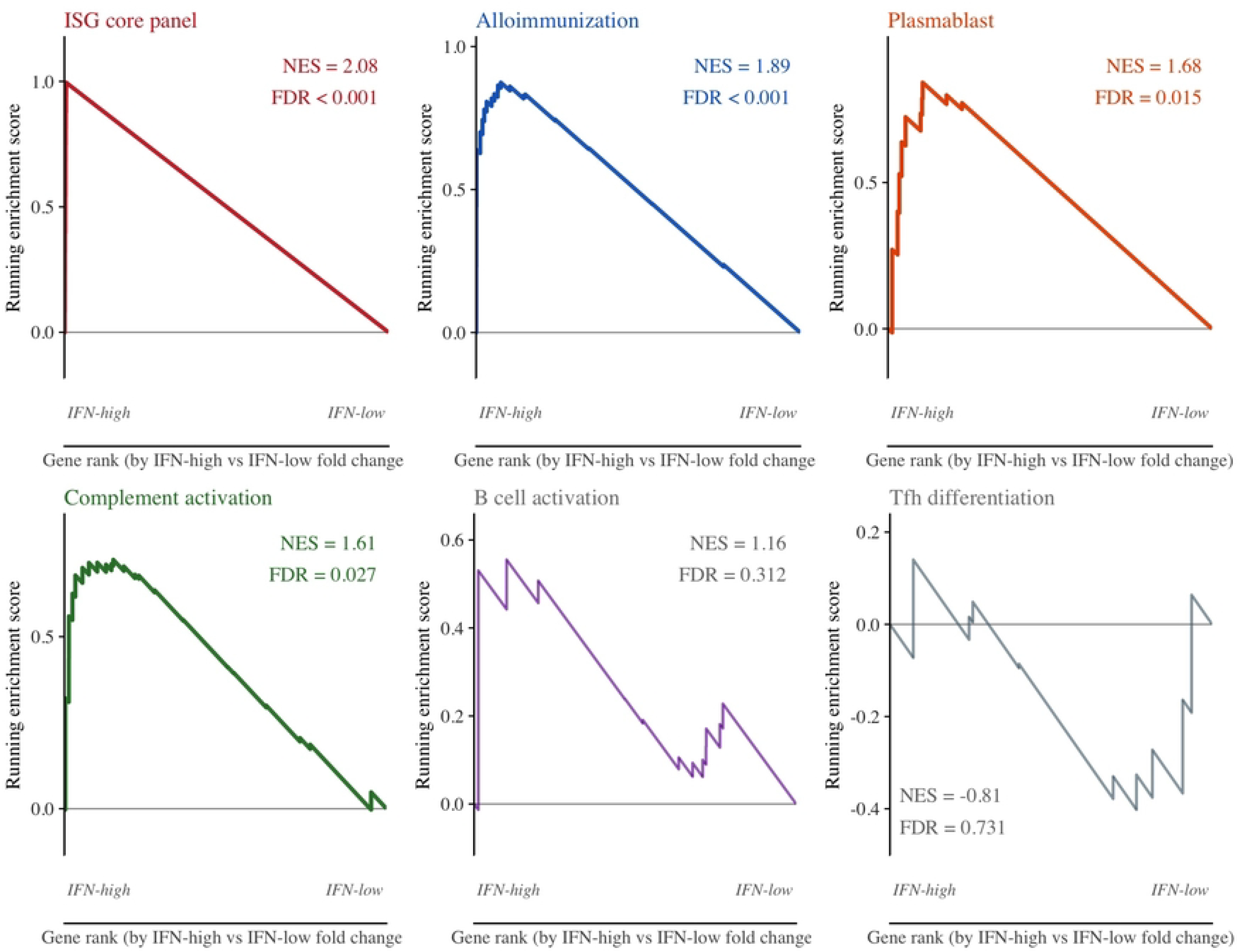
Gene set enrichment analysis (GSEA) enrichment plots for six gene sets in IFN-high versus IFN-low SLE patients (GSE72509 discovery cohort). Each panel shows the running enrichment score curve (top) and gene hit positions along the ranked list (rug plot, bottom). Genes were ranked by sign(log_2_FC) × [−log_10_(FDR)]. Normalized enrichment score (NES) and FDR q-value are shown in each panel. Significant enrichment (FDR q < 0.05) was observed for three gene sets: ISG core panel (NES = +2.70, FDR < 0.001), alloimmunization-associated (NES = +2.34, FDR < 0.001), and plasmablast differentiation (NES = +1.94, FDR = 0.002). Complement activation showed a trend toward enrichment that did not reach significance after FDR correction (NES = +1.52, FDR = 0.066). B cell activation (NES = +0.82, FDR = 0.691) and Tfh differentiation (NES = −1.09, FDR = 0.334) were not significantly enriched. NES, normalized enrichment score; FDR, false discovery rate (Benjamini–Hochberg correction across the six gene sets tested). 1,000 permutations; seed = 42.

### Alloimmunization Signature Correlates with IFN-I Score Across All Three Independent Cohorts

Spearman correlation between continuous IFN-I scores and pathway module scores was computed independently in each cohort (Figure 5). In the discovery cohort (GSE72509, n = 99 SLE), the alloimmunization signature showed the strongest association (ρ = +0.77, p < 0.001), followed by complement (ρ = +0.56, p < 0.001) and plasmablast differentiation (ρ = +0.39, p < 0.001). Tfh showed no association (ρ = −0.158, ns).

**Figure 5.**
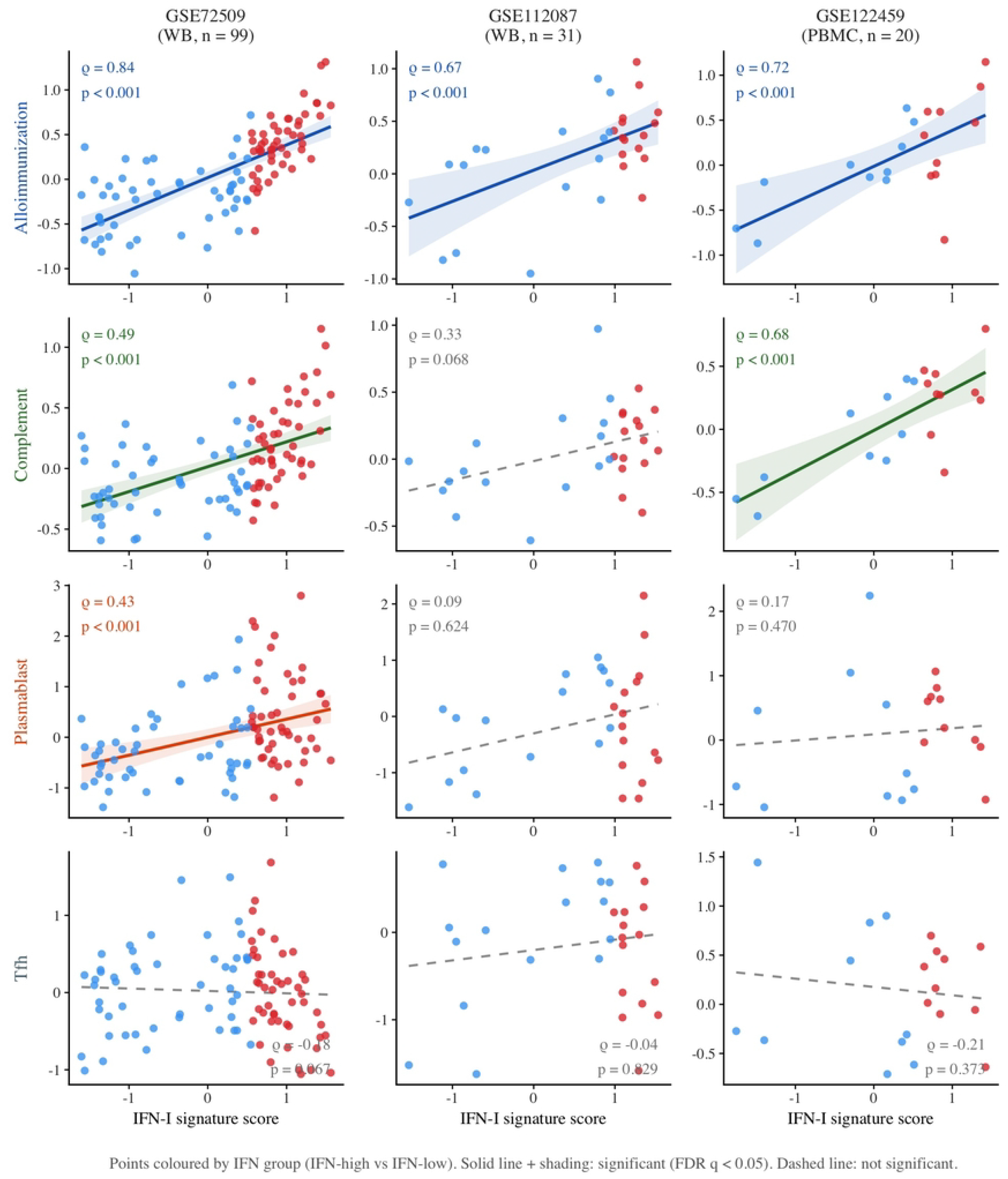
Spearman correlations between IFN-I signature score and pathway module scores across three independent cohorts (SLE patients only). Each panel shows individual SLE patient data points (IFN-I score on x-axis; pathway module score on y-axis). Regression lines are shown for significant correlations (FDR q < 0.05; solid line) and omitted for non-significant associations (dashed line). Shaded area indicates 95% bootstrap confidence interval. Spearman ρ coefficient and nominal p-value are shown in each panel. Rows: alloimmunization signature, complement activation, plasmablast differentiation, Tfh pathway (top to bottom). Columns: GSE72509 circles (n = 99), GSE112087 squares (n = 31), GSE122459 diamonds (n = 20), left to right.

In the replication cohort (GSE112087, n = 31 SLE), the alloimmunization correlation was replicated (ρ = +0.51, p = 0.003, FDR q = 0.022). Plasmablast differentiation did not show a significant association in this cohort (ρ = +0.16, ns). Complement showed a nominally significant positive correlation (ρ = +0.40, p = 0.025) but did not survive FDR correction (q = 0.087), directionally consistent with the other cohorts; the failure to reach FDR significance is likely attributable to the smaller sample size (n = 31). Tfh showed no significant association in this cohort (ρ = −0.052, ns). In the PBMC validation cohort (GSE122459, n = 20 SLE), alloimmunization showed strong correlation (ρ = +0.60, p = 0.005, FDR q = 0.025) and complement showed significant positive correlation (ρ = +0.58, p = 0.007, FDR q = 0.025), providing cross-platform replication. Plasmablast showed a positive but non-significant trend (ρ = +0.075, ns), consistent with limited statistical power at n = 20.

Across all three cohorts, the alloimmunization signature was the most consistently significant pathway, reaching significance in every independent dataset (nominal p < 0.05 and FDR q < 0.05 in all three). Plasmablast differentiation was significant in one of three cohorts (GSE72509 only; FDR q < 0.001). Complement was nominally significant (p < 0.05) in all three cohorts but survived FDR correction in two of three (GSE72509: q < 0.001; GSE122459: q = 0.025); the association did not survive FDR correction in GSE112087 (p = 0.025, q = 0.087). Tfh differentiation showed no association in any cohort. Gene set member counts differ between the GSEA analysis (genes expressed above detection threshold in GSE72509 after filtering) and the Spearman analysis (all annotated genes available per dataset); this difference is expected given dataset-specific expression filtering thresholds. As internal specificity controls, T cell generic module scores showed a negative correlation with IFN-I score in the discovery cohort (ρ = −0.266, p = 0.008) but did not reach significance in the replication (ρ = −0.153, ns) or PBMC validation (ρ = −0.415, p = 0.069) cohorts. NK cell scores showed no consistent association (GSE72509: ρ = −0.073, ns; GSE112087: ρ = −0.162, ns; GSE122459: ρ = +0.400, ns).

## Discussion

In this multi-cohort transcriptomic study, we provide what is, to our knowledge, the first human transcriptomic evidence that IFN-I pathway activity in SLE is associated with a pro-alloimmunization immune landscape, characterized by consistently elevated alloimmunization-associated gene expression with directional evidence supporting complement activation and plasmablast differentiation that warrants further validation. The alloimmunization signature correlation was replicated across all three independent cohorts, two using whole blood RNA-seq and one PBMC RNA-seq. This cross-compartment replication is biologically informative beyond technical concordance: persistence of the alloimmunization association in PBMC establishes that the core signal resides in mononuclear cells and is not dependent on granulocyte or erythrocyte contributions unique to whole blood. IFN-I-based stratification of SLE patients into biologically distinct subgroups has been previously validated as a clinically meaningful approach to capturing disease heterogeneity^14^; our findings extend this framework by linking IFN-I pathway strength specifically to transcriptomic programs governing alloimmunization risk.

Our findings provide human transcriptomic support for the mechanistic framework established by Gibb and colleagues^2,15^. Their work showed that IFNAR signaling is required for efficient RBC alloimmunization, that Nrf2 deficiency promotes alloimmunization by unleashing IFN-I responses to erythrophagocytosis, and that Nrf2 activation suppresses IFN-I and reduces alloimmunization in pre-clinical transfusion models^15^. IFN-high SLE patients exhibit the gene expression features predicted by the murine model; whether this transcriptomic pattern predicts actual alloimmunization outcomes requires prospective study.

A potential concern is whether the correlation reflects mathematical overlap between the IFN-I scoring panel and the alloimmunization gene set. These are distinct: none of the 14 IFN-I scoring genes appear among the 20 alloimmunization genes, ruling out direct instrument overlap. A separate but related question is whether some alloimmunization genes are transcriptionally regulated by IFN-I—and 8 of the 20 are, in the sense that their expression is induced by IFN-I signalling. However, being IFN-regulated is not the same as being an IFN scoring gene: the 14-gene scoring panel consists of canonical interferon-stimulated genes used as a measurement instrument, whereas the 8 IFN-regulated alloimmunization genes are downstream effector genes (innate nucleic acid sensors and IFN regulatory factors) that IFN-I induces as part of its broader immune programme. Excluding these 8 and retaining only the 12 non-IFN-adjacent alloimmunization genes (Fcγ receptors, complement components, and plasma cell markers) still yielded a robust correlation with IFN-I score in the discovery cohort (ρ = +0.61, p = 2.1 × 10^−11^; Figure 5), representing 79% of the full-set effect and remaining directionally consistent in both replication cohorts. This sensitivity analysis argues against the association being an artefact of shared transcriptional regulation; the correlation persists even in genes with no functional connection to IFN-I signalling.

Complement activation correlated significantly with IFN-I score in the discovery and PBMC validation cohorts (Figure 5), with borderline GSEA enrichment in the discovery cohort (FDR = 0.066; Figure 4), which we interpret as exploratory. The compartment-dependent pattern—reaching significance in the smaller PBMC cohort (q = 0.025) but not the larger whole-blood cohort (q = 0.087)—is consistent with a monocyte-driven signal, as C1Q complex components are predominantly expressed in monocytes and diluted in whole blood by granulocytes. Complement is constitutively dysregulated in SLE^18^, and IFN-I transcriptionally upregulates C1q components, providing a plausible mechanistic link to alloantigen opsonization. Plasmablast differentiation was significant in the discovery cohort but unreplicated in either replication cohort, including the PBMC compartment where B-lineage cells are enriched; this argues for an intrinsically weaker association that may require larger cohorts to detect consistently. IFN-I promotes plasmablast formation via STAT1 and STAT3 signaling^19^. Tfh pathway scores were non-significant across all three cohorts, consistent with Tfh-independent mechanisms of alloimmunization in the murine literature.

### Clinical Implications and Therapeutic Relevance

The IFN-I transcriptomic signature is already used in clinical practice as a stratification tool in SLE; our findings suggest an additional utility as a pre-transfusion risk indicator. SLE patients with high IFN-I scores may warrant heightened alloimmunization surveillance and extended antigen matching. Anifrolumab, a monoclonal antibody blocking IFNAR approved for active SLE^16^, reduces the IFN-I transcriptomic signature and may also reduce alloantibody formation in transfused SLE patients—a testable hypothesis with meaningful clinical implications^17^. Prospective transfusion cohort studies in SLE patients stratified by IFN-I score and anti-IFN therapy status are needed to evaluate these possibilities.

Several limitations warrant acknowledgment. First, all cohorts are cross-sectional without prospective alloimmunization outcome data, precluding direct assessment of whether IFN-I score predicts subsequent alloantibody formation. Second, bulk RNA-seq cannot resolve cell-type-specific contributions; however, partial Spearman correlations controlling for T cell and NK cell module scores left the alloimmunization and complement associations virtually unchanged, arguing against cell-composition confounding. Third, the alloimmunization gene set has not been prospectively validated as a clinical predictor. Finally, cohort sizes across all three datasets limit statistical power for secondary pathway analyses, particularly for complement and plasmablast pathways where effect sizes appear smaller.

In conclusion, this multi-cohort transcriptomic study provides the first human-level evidence that IFN-I pathway strength in SLE is coupled to a broad alloimmunization-prone immune landscape. The consistency of the alloimmunization association across three independent cohorts, two sample types, and a pre-specified analytical design strengthens confidence in the finding. Whether IFN-I score predicts actual alloantibody formation in transfused SLE patients, and whether targeting the IFN-I pathway with approved therapies modulates alloimmunization risk, are open questions that require prospective evaluation with transfusion endpoints.

## Data Availability

No data was generated by this study. The following existing data sources were used: GSE72509 from NCBI Gene Expression Omnibus available via https://www.ncbi.nlm.nih.gov/geo/query/acc.cgi?acc=GSE72509 GSE112087 available via https://www.ncbi.nlm.nih.gov/geo/query/acc.cgi?acc=GSE112087 GSE122459 available via https://www.ncbi.nlm.nih.gov/geo/query/acc.cgi?acc=GSE122459.

## Data Availability Statement

All data are publicly available from NCBI Gene Expression Omnibus (GEO): GSE72509 (https://www.ncbi.nlm.nih.gov/geo/query/acc.cgi?acc=GSE72509), GSE112087 (https://www.ncbi.nlm.nih.gov/geo/query/acc.cgi?acc=GSE112087), and GSE122459 (https://www.ncbi.nlm.nih.gov/geo/query/acc.cgi?acc=GSE122459). Analysis code is available upon reasonable request to the corresponding author.

## Acknowledgments

The authors thank the original investigators who deposited the datasets GSE72509, GSE112087, and GSE122459 to NCBI GEO and made them publicly available.

This research received no specific grant from any funding agency in the public, commercial, or not-for-profit sectors.

## Author contributions

**JY: Conceptualization, Data Curation, Formal Analysis, Visualization, Methodology, Supervision, Writing – Original Draft, Writing – Review & Editing**.

## Competing Interests

The authors declare no competing interests.

## Notes

### Competing Interest Statement

The authors have declared no competing interest.

### Funding Statement

The author(s) received no specific funding for this work.

### Author Declarations

This study is a computational re-analysis of publicly available, de-identified transcriptomic data deposited in NCBI Gene Expression Omnibus (GSE72509, GSE112087, GSE122459). No new human participants, specimens, or identifiable data were collected. Institutional ethics approval was not required.

